# Obesity and Smoking: A Tale of 2 Risk Factors with Implications for the Next Pandemic

**DOI:** 10.1101/2023.03.23.23287630

**Authors:** Mary L. Adams

## Abstract

**Background:** In 1990, two risk factors that would figure prominently in the COVID-19 pandemic were on divergent paths in the US. The smoking rate was 23.5% and dropped to 13.5% in 2021, while the obesity rate was 11.5% and increased 186% to 33.0%.

**Objective:** The study objective was to compare the global impact of those risk factors on COVID deaths to help prepare the US for future pandemics.

**Methods:** Stata and Excel were used to regress global COVID deaths on obesity and smoking before and after vaccines were available, and US deaths/day were compared pre-and post-vaccines.

**Results:** Obesity was associated with global COVID deaths, with R^2^ as high as 0.87 for cumulative data with slightly lower R^2^ and coefficients for post-vaccines. For 9 regressions of deaths on obesity, all P values (overall and coefficients) were <0.05 while for regressions on smoking, no P values were < 0.05. Of the 1.1 million US deaths, the death rate/day post-vaccines was 59% of that pre-vaccines. If the US obesity rate had remained 11.5%, estimates suggest 800,000+ lives could have been saved. US smoking rate was reduced 42% by multiple strategies using support from a 1998 multi-billion-dollar settlement between states and tobacco companies.

**Conclusion:** Vaccines have limited ability to reduce total COVID deaths, with obesity remaining a key factor in death rates. Results suggest that lower obesity rates are needed to further reduce US COVID deaths, potentially saving thousands of lives in future pandemics. Lessons from reducing smoking rates might prove useful.

## Background

When COVID-19 first appeared in China early results from there showed that cardiovascular disease, diabetes, chronic respiratory disease, hypertension, and cancer increased the case fatality rate (1). Based on those results, 45% of all US adults were estimated to have conditions that increased rates of hospitalizations and death (2). When US hospitalization data were used and obesity was added to the list the percentage at risk increased to 56% (3). Three years later, as of January 27, 2023, 1.1 million US deaths have been attributed to the pandemic (4) making COVID-19 the 3^rd^ leading cause of death in 2020 when the death toll stood at 350,831 (5). Over half of US COVID deaths occurred after effective vaccines became available in early 2021 and the death toll is still climbing.

The World Obesity Federation (WOF) noted early in the pandemic (6) that overweight was a critical factor in COVID deaths. In countries where the prevalence of overweight was >50%, the death rate from COVID was about 10 times that in countries where overweight was <50%, with a steep increase in death rates above 50%. A global study of COVID deaths (7) and obesity pre-vaccines found that as much as 90% of the variation in death rates among 167 countries could be explained by differences in obesity rates alone. Comparing that study with the WOF results (6) suggests that an obesity rate of 15% corresponds to an overweight rate of 50%, with death rates ranging from < 1/million persons up to 1,892/million persons pre-vaccines (2/25/2021) for obesity rates between 2.1% and 37.9% (7). With the second highest obesity rate in the world at 36.2%, the US had one of the highest death rates and the most deaths of any country. For the 57 countries with obesity rate <15%, the average death rate was 40.8/million making the US death rate of 1,562/million persons at that time 38.3 times the average death rate in countries with obesity rates <15%.

Noting that as recently as 1990 the US obesity rate was 11.5% (8, 9), it seemed natural to wonder how the pandemic consequences might have been different if the obesity rate was still 11.5%. Or alternatively, how many of those 1.1 million US lives might have been saved with *any* lower obesity rate? Also noting that the smoking rate in 1990 was 23.5% and is now down to 13.5%, a brief look at how that was done might suggest strategies that could be used to lower the US obesity rate.

The objective for this study was to compare the impact of smoking and obesity (BMI ≥30) on COVID and propose strategies that might reduce deaths in future pandemics. While the impact of COVID on the US is the focus, most of the analysis will use global data because a wide range of obesity rates was found to be needed for such analysis (7). Two hypotheses will be tested: 1) Is there still a linear regression model for COVID deaths and obesity after vaccines (and treatments & natural immunity) became available as was shown globally for the period pre-vaccines (6, 7); 2) Is there a similar linear association between COVID deaths and current smoking over the course of the pandemic. Depending on results from hypothesis testing estimates could be made of potential death rates resulting from successful reduction of risk factor rates at various levels. Results should help to put the impact of smoking and obesity on US COVID deaths into perspective and suggest strategies that might be used to reduce deaths in future pandemics.

## Methods

All data are publicly available and referenced. For global data, an Excel file used previously (7) with deaths as of February 25, 2021 and 2016 obesity data (10) was amended to include 2020 smoking rates (11). For global data post-vaccines, death rates as of 2/27/2023 (3) were used with obesity rates from 2016 (10). The difference between death rates in each country between the 2 dates was used for the 2 years post-vaccines (2/25/2021-2/27/2023) and to determine simple proportions. Data were sorted by country population as done in that previous study (7) and analysis was done on groups of 10, starting with the 10 most populous countries (China, India, USA, Indonesia, Pakistan, Nigeria, Brazil, Bangladesh, Russia, and Mexico, with 59.1% of total world population) up to 60 countries with 91.6% of total), and then all 120 with data on both deaths and obesity. Stata version 14.1 (Stata Corp LP, College Station, TX) was used for regression which produced coefficients, Y intercepts, R^2^ and adjusted R^2^ plus t tests of each coefficient and overall F tests. Regression in Excel produced the graphs. Data points for US red and blue states (based on voting in the 2016 Presidential election) from earlier in February 2023 (12) were added to the graphs of death rates using the obesity rate in 2019 for the respective state groups. Results for the ten countries listed were used to obtain estimates of COVID deaths for different obesity rates. As reported in that earlier study (7), only using a very wide range of obesity rates such as obtained globally will result in data that can be used for estimates.

## Results

Global obesity rates were associated with COVID deaths as shown in Figures 1 and 2 for the ten most populous countries, cumulative and post vaccine results only. R^2^ values (unadjusted) were 0.867 for the cumulative 3 years and 0.71 for the post vaccine period of 2 years and slopes (coefficients) were 116 and 65 respectively. All the coefficients for models with only obesity had P values for t-tests and for overall F tests <0.05 while none of the models with only smoking had any P values <0.05 for those tests. The red and blue dots represent the values for red and blue US states at a slightly earlier time in February 2023 and use actual obesity rates from a different source (12) and are for perspective. Results of regression analysis for obesity and smoking are summarized in Table 1. Estimates made using the formulas from the cumulative data have large margins of error but suggest that between 800,000 and 869,000 deaths could have potentially been saved over the course of the pandemic if the US obesity rate was still 11.5% and from 300,000-680,000 lives could be saved by reducing obesity rates to 20% - 25%.

**Table 1.**
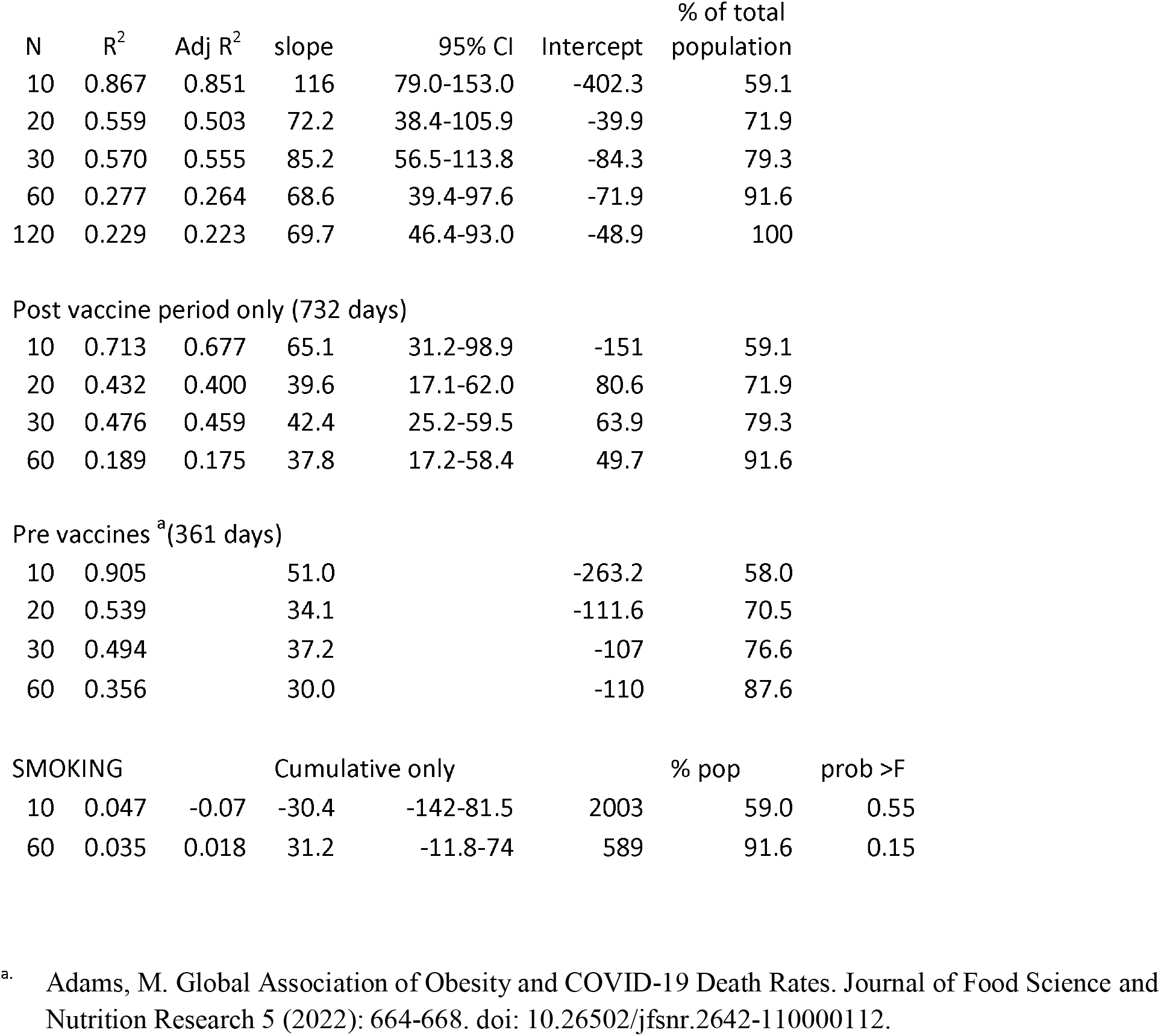
Regression results from Stata Regression of COVID deaths to 2/27/2023 on obesity (1093 days)

| N | R<sup>2</sup> | Adj R<sup>2</sup> | slope | 95% CI | Intercept | % of total population |
| --- | --- | --- | --- | --- | --- | --- |
| 10 | 0.867 | 0.851 | 116 | 79.0-153.0 | -402.3 | 59.1 |
| 20 | 0.559 | 0.503 | 72.2 | 38.4-105.9 | -39.9 | 71.9 |
| 30 | 0.570 | 0.555 | 85.2 | 56.5-113.8 | -84.3 | 79.3 |
| 60 | 0.277 | 0.264 | 68.6 | 39.4-97.6 | -71.9 | 91.6 |
| 120 | 0.229 | 0.223 | 69.7 | 46.4-93.0 | -48.9 | 100 |
| Post vaccine period only (732 days) |  |  |  |  |  |  |
| 10 | 0.713 | 0.677 | 65.1 | 31.2-98.9 | -151 | 59.1 |
| 20 | 0.432 | 0.400 | 39.6 | 17.1-62.0 | 80.6 | 71.9 |
| 30 | 0.476 | 0.459 | 42.4 | 25.2-59.5 | 63.9 | 79.3 |
| 60 | 0.189 | 0.175 | 37.8 | 17.2-58.4 | 49.7 | 91.6 |
| Pre vaccines <sup>a</sup>(361 days) |  |  |  |  |  |  |
| 10 | 0.905 |  | 51.0 |  | -263.2 | 58.0 |
| 20 | 0.539 |  | 34.1 |  | -111.6 | 70.5 |
| 30 | 0.494 |  | 37.2 |  | -107 | 76.6 |
| 60 | 0.356 |  | 30.0 |  | -110 | 87.6 |

| SMOKING |  |  | Cumulative only |  |  | % pop | prob >F |
| --- | --- | --- | --- | --- | --- | --- | --- |
| 10 | 0.047 | -0.07 | -30.4 | -142-81.5 | 2003 | 59.0 | 0.55 |
| 60 | 0.035 | 0.018 | 31.2 | -11.8-74 | 589 | 91.6 | 0.15 |

<sup>a</sup>. Adams, M. Global Association of Obesity and COVID-19 Death Rates. Journal of Food Science and Nutrition Research 5 (2022): 664-668. doi: 10.26502/jfsnr.2642-110000112.

**Table 2.** Estimates of deaths/million expected at different obesity rates Actual lives lost in US per million to 2/27/2023=3,421

| Target obesity rate | 11.5 | 20 | 25 |
| --- | --- | --- | --- |
| Using formula for 10 countries | 931.7 | 1917.7 | 2497.7 |
| Fraction of actual | 0.27 | 0.56 | 0.73 |
| Estimated lives saved | 800,418 | 483,376 | 296,881 |
| Using formula for 30 | 895.5 | 1619.7 | 2045.7 |
| Fraction of actual | 0.26 | 0.47 | 0.60 |
| Estimated lives saved | 812,058 | 579,196 | 442,219 |
| Using formula for 60 | 717 | 1300.1 | 1643.1 |
| Fraction of actual | 0.21 | 0.38 | 0.48 |
| Estimated lives saved | 869,453 | 681,961 | 571,672 |

**Figure 1.**
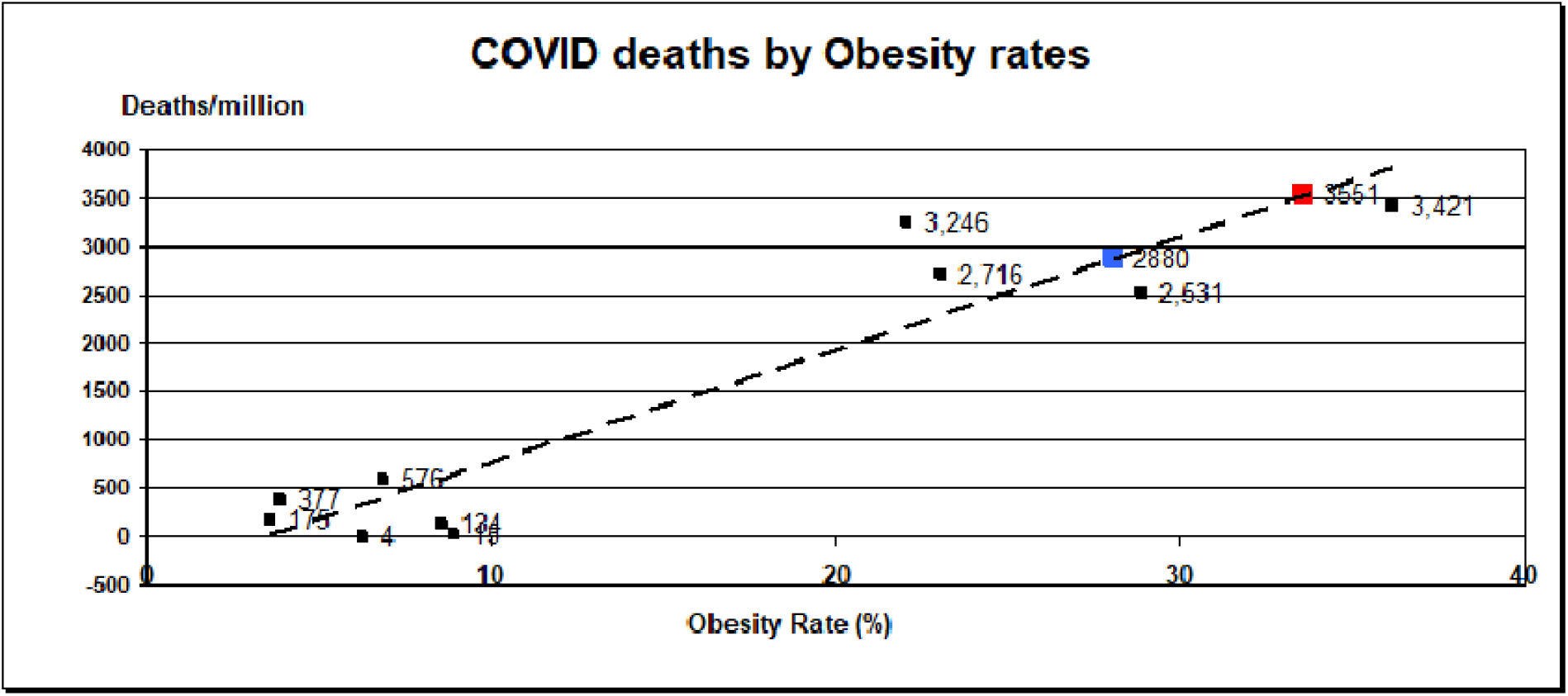
Graph of COVID-19 deaths through 2/27/2023 by 2016 obesity rates for the 10 most populous countries with data for both. The 3,421 data point is for the US and the blue and red data points represent the positions of the actual data points for blue and red states respectively (assigned based on how the state voted in 2016 Presidential election) at a slightly earlier February 2023 date. Dotted line is fitted linear trend line (least squared fit).

**Figure 2.**
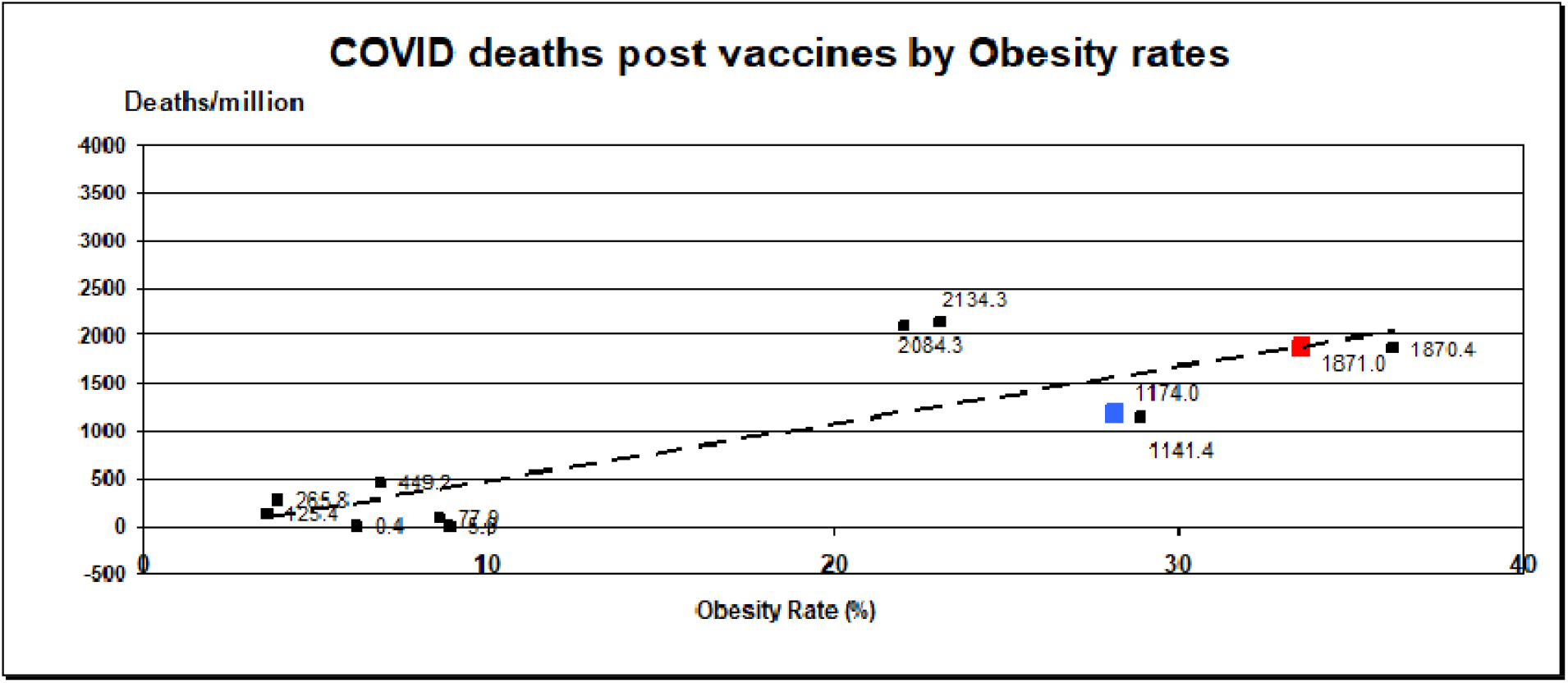
Graph of COVID-19 deaths between 2/25/2021 and 2/27/2023 by 2016 obesity rates for the 10 most populous countries with data for both. The 1,870 data point is for the US and the blue and red data points represent the positions of the actual data points for blue and red states respectively (assigned based on how the state voted in 2016 Presidential election) at a slightly earlier February 2023 date. Dotted line is fitted linear trend line (least squared fit).

The total number of COVID-19 deaths worldwide was 6.798 million as of February 27, 2023, which included 2.513 million deaths as of 2/25/2021, before vaccines. Thus, the difference between those figures of 4.285 million represents the number of pandemic deaths that were recorded in the 2 years when vaccines were assumed to be available which is 63% of total pandemic deaths to 2/27/2023. Using a different data source (12) and slightly different dates as noted in methods, this fraction varied between the red and blue states deaths in the post vaccine period (since 4/19/2021) and represent 40.6% of the total pandemic deaths in blue states and 52.7% in red states. Figures 1 and 2 each include those data points which show the difference in obesity rates between red and blue states. The vaccination rate in blue states (12) was about 10 percentage points higher than that in red states. Calculating these fractions in terms of deaths per day results in different figures. In the US this fraction was calculated from study data as post vaccine deaths/day as a percentage of cumulative deaths/day and was 81.6%; calculated as the post-vaccine death rate/day as a fraction of the pre-vaccine rate/day the result was 59%. For the global data, these fractions are 94% and 84% respectively, and expected to have great variation between different countries.

## Discussion

Several observations should be clear from the results, starting with the graphs. Obesity matters to COVID death rates, even when effective vaccines are available. Results in Figures 1 & 2 indicate that nearly 90% of the difference in cumulative COVID death rates between these 10 countries can be explained by obesity alone and this only drops to about 70% when limited to the time when vaccines were available. This latter R^2^ value does not leave a lot of room for the contribution of other factors, including vaccines and treatments. The graphs make it quite clear that the high COVID death rate in the US is associated with its high obesity rate and almost any reduction in that obesity rate should save lives. Vaccines are especially effective on an individual basis (13) but they do not seem able to reduce the death rate appreciably in countries where the obesity rate is high. On the other hand, these results indicate that current smoking is not associated with COVID deaths. It is still possible that lifetime smoking or results of smoking such as chronic lung disease would be associated, but that was not the goal of this paper. In looking for ways to save lives in the next pandemic it seems that obesity might be a good place to start. Obesity has already been shown to be associated with COVID deaths early in the pandemic (6,7) but these new results indicate that obesity still matters while vaccines are available.

Regression results in Table 1 indicate that adding more countries to regression results lowers the R^2^ value as it adds more countries that have death rates that are outliers. It also appears that the slope for the post-vaccine period is not as steep as that for the cumulative data, as would be expected, but still reflects an association between obesity and deaths. On a per day basis, the post-vaccine global death rate per day was 84% of the pre-vaccine daily death rate and 94% of the cumulative death rate/day which should be a very rough estimate of the global effect of vaccines. It is also likely that this rate varies widely from country to country but overall, suggests that the impact of vaccines only slightly reduced the COVID death rate from the time before vaccines were available. In the US those same rates per day at 59% and 81.6% respectively, were much lower than the global rates and appear to suggest perhaps vaccines had more of an impact in the US than other countries. The US reduction in deaths per day between post- and pre-vaccines represents a 41% reduction in death rate that might be attributed to vaccines, treatments, and/or natural immunity. But the simple math showing that 63% of all COVID deaths in the world occurred after vaccines started to become available should be evidence that COVID-19 is not totally over.

It is clear in every graph that countries with obesity rates <15% have VERY low death rates. It is also likely that the reason there are not more papers reporting associations between obesity and COVID deaths is because a very large difference in obesity rates is needed to detect the association (7). Not apparent from the graph is that the six countries with obesity rates <15% are all in Africa or Asia. Expanding to more countries does not change that finding – all 57 countries with obesity rates <15% are in Africa or Asia (7) and have very low death rates. Because there are over 50 countries with such low obesity rates and matching low death rates it seems highly unlikely that they are all reporting unreliable death data. As noted, those countries with very low obesity rates are critical to this analysis (7). The graphs with only 10 countries look almost too good to be true, but they are actual data. It appears to be a coincidence that none of those 10 countries have large deviations from the fitted line – and that includes the US.

### Encouraging findings

Figures 1 and 2 also include data points from red and blue states in the US at a slightly earlier date in February 2023 (12) showing that the blue states have lower obesity rates and death rates than red states. In Figure 1 (cumulative results) those US data points appear to be right on the fitted line while the blue point is below the line for the post-vaccine data. The location of the points could reflect only the differences in obesity rates or vaccination rates, or both. The red and blue points appear to be consistent with the different obesity rates among the states: 28.1% for blue states and 33.6% for red states but it could also reflect the approximately 10% higher vaccination rates in blue states (12), leading to lower deaths. Unlike the estimates that predicted 800,000 or more lives could have been saved if the US obesity rate was still 11.5%, these are actual data points representing the difference in death rates between red and blue states. The finding that these data points fall on the fitted least squares line for the regression results suggests that even if they are not precise they might represent death rates differences corresponding to relatively small differences in obesity. In a previous US study (12) post vaccines, the impact of vaccines on deaths rates was found to be the greatest of three factors in the model, none of which was obesity alone. That study (12) found that from 4/19/2021-2/28/2022 the number of deaths per day in blue states dropped 36.6% while deaths per day rose 9.4% in red states. That seems to suggest that vaccines also play a role in saving lives, along with obesity. Thus, these differences might reflect more than differences in obesity rates but should help to show achievable improvements in death rates.

Other studies have included factors in addition in obesity in regression models that might suggest other ways to save lives in the next pandemic. One such study (14) of 30 countries included population density, the age structure of the population, population health, gross domestic product (GDP), ethnic diversity, and how the pandemic was handled. That model explained 63% of the difference in death rates between countries. That result is not inconsistent with the results in this paper where obesity was found to explain up to 87% of the difference in death dates between countries depending on number of countries included. Based on the list of factors included in that model (14), obesity and how the pandemic were handled appear to be the only potentially modifiable factors that could save lives in a future pandemic.

Results from other studies may help explain why obesity apparently remains such a key factor in COVID deaths even when vaccines are available. Not only does obesity increase the risk of death from COVID-19 (3), but obesity can lead to chronic inflammation and impaired immune response which can increase the risk of developing COVID and severe illness from it (15-17). There is also evidence that vaccines are less effective in obese individuals compared with those who are not obese (15). These same factors might also contribute to a higher rate of breakthrough infections in fully vaccinated people (18). These and other unknown influences may affect the association of obesity with COVID death rates, but nothing appears to change the key result that obesity is linked to an increase in COVID deaths. These results seem to reinforce the idea that the best strategy for reducing deaths from future pandemics will involve reducing obesity rates. The results from studies showing obesity affects immunity (15-17) and vaccines (15, 18) suggest that a study of the effect of obesity on COVID transmission (i.e. with cases as outcome) might be useful, bearing mind the probable need for the inclusion of a wide range of obesity rates.

This brings the discussion back to the second of the 2 risk factors. While reducing smoking would appear to have little or no impact on COVID death rates, perhaps the strategies that were used between 1990 and 2021 to reduce the US smoking rate by 42% could provide inspiration. A 42% reduction in the US obesity rate used in the global analysis in this study would bring the US obesity rate down to between 20-25% and save an estimated 300,000-680,000 deaths compared with current COVID-19 deaths. A key factor in that huge reduction in US smoking rates appears to be the Master Settlement Agreement (MSA) between tobacco companies and most states (19) finalized in 1998. That process began in 1994 when a few states sued major tobacco companies to recover Medicaid and other health care costs they incurred to treat illnesses caused by smoking. After 4 states settled in 1997, more states joined until the Master Settlement that involved most states and tobacco companies. The agreement included annual payments to states starting in 2000, which continue in perpetuity, but the states were not restricted in how the funds could be spent. Payments to the states through July 2018 totaled $126 billion but <1% was earmarked for state tobacco prevention programs. The basis of the MSA, that the *products the company made were harming the health of citizens which was costing the states money*, appears key. Included in the MSA were severe restrictions on advertising, prohibited lobbying against specific legislation and activities that would seek to hide adverse effects of tobacco. Increased taxation on tobacco products was also used. Perhaps its greatest impact was just the recognition it brought to the negative effects of tobacco and the link to health care costs. The costs of obesity should also be considered in this case: estimates have put the total cost of obesity in the US between $172.7 billion (20) and $260.6 billion (21) using different methods. These costs amount to between $1,800 (20) and $2,500 (21) in extra costs for each obese adult. Those estimates were made pre-pandemic and the pandemic itself was estimated to cost the US $16 trillion (22) through the fall of 2021.

With or without something akin to a MSA, there are steps that could be taken to reduce obesity. One would be taxation of specific products such as sugar sweetened beverages that have been shown to be associated with higher obesity rates (6). Two simple behaviors appear to help in reducing obesity: eating fruits and vegetables 5 or more times a day and getting 150 minutes or more of moderate exercise a week, such as walking. Recent US survey data (8) have shown that the 8.2% of adults who engage in both behaviors had an obesity rate of 24.7% compared with the 47% who did neither and had an obesity rate of 36.8%. Reporting both behaviors was also associated with lower rates of hypertension, heart disease, diabetes, and COVID deaths. Again, these are actual data and not estimates. However, estimates indicate as noted above, that an obesity rate between 20-25% could potentially save an estimated 300,000-680,000 deaths compared with actual deaths from COVID-19. This appears to be an easy change to encourage people to make with potentially large health benefits.

### Limitations

A reminder that these results are for countries (or states) and may not apply to individuals. For example, smoking may increase the risk of complications and death for persons who smoke but not show an association when the measure is current smoking rates in different countries. Data show clearly that while obesity is a factor in COVID death rates, factors other than obesity are involved in some countries that either increase or decrease the death rates. The quality of the data is unlikely to be uniform and in the later stages of the pandemic, the dates when data were available are not always consistent. For example, the US data points (for red and blue states) were not obtained on the exact same dates as the global data. It appears that death data are not updated as consistently now as they were earlier in the pandemic, so it is more difficult to match dates. An extra time point post-vaccines would have been helpful.

### Conclusion

These results clearly show that the availability of vaccines doesn’t eliminate pandemic deaths and that obesity remains a key factor in the 1.1 million US COVID deaths to date. Monetary costs of obesity were high even before the pandemic which undoubtedly increased those costs immensely. The World Obesity Federation (6) believes that obesity is likely to be a factor in any future pandemics. The estimates suggesting thousands of deaths could have been saved in the US with a much lower obesity rate and the actual differences in death rates between red and blue states with relatively small differences in obesity rates, suggest that it should be well worth any attempts to reduce obesity rates. Obesity rates for US adults who eat fruits and vegetables ≥ 5 times a day and engage in moderate exercise are also encouraging. Results from other studies do not seem to offer any viable alternatives for strategies to reduce pandemic deaths.

## Data Availability

All data are available online at links provided in references.

https://www.worldometers.info/coronavirus/

https://ourworldindata.org/smoking#prevalence-of-smoking-across-the-world

https://web.archive.org/web/20200630202020/

https://www.cia.gov/the-world-factbook/rankorder/2228rank.html

## No funding was received for this study

The author declares no conflicts of interest.

